# Short-term effects of “Yijinjing Wohu Pushi” posture-voice therapy on speech in Parkinson’s disease after STN-DBS

**DOI:** 10.1101/2023.12.13.23299527

**Authors:** Xin Sun, Mei Yang, Jin Yan, Linbin Wang, Yuxin Sun, Yong Wang, Shiqing Yan, Dianyou Li, Chuanxin M. Niu

**Affiliations:** Department of Rehabilitation Medicine, Ruijin Hospital, Shanghai Jiao Tong University School of Medicine, Shanghai, China, 200025; School of Medicine, Shanghai Jiao Tong University, Shanghai, China, 200025; The Affiliated Chuzhou Hospital of Anhui Medical University, Anhui, China, 239000; Institute of Science and Technology for Brain-Inspired Intelligence (ISTBI), Fudan University, Shanghai, China, 200433; Department of Traditional Chinese Medicine, Ruijin Hospital, Shanghai Jiao Tong University School of Medicine, Shanghai, China, 200025; Shanghai Institute of Inheritance and Daoyin Medicine, Shanghai, China, 200437; Department of Neurosurgery, Center for Functional Neurosurgery, Ruijin Hospital, Shanghai Jiao Tong University School of Medicine, Shanghai, China, 200025

**Keywords:** Parkinson’s disease, rehabilitation, vocalization, posture, deep brain stimulation

## Abstract

Hypokinetic dysarthria in Parkinson’s Disease (HD-PD) deteriorates patients’ quality of life by impeding communication and social engagement. Existing treatments like levodopa drugs and deep brain stimulation (DBS) can improve motor symptoms but fall short in addressing speech-related impairments; effective speech therapies tend not to mandate the posture, which potentially hinders vocal performance. Here we first proposed a therapeutic protocol that specified a lunging-and-clawing posture (Wohu Pushi method from Yijinjing) during vocalization training. The protocol aimed to ensure that PD patients could accomplish the training even with moderate motor impairments. A total of 20 HD-PD patients with implanted stimulators in subthalamic nuclei (STN-DBS) participated in one session of training, either following our posture-voice therapy (PVT) or the conventional voice therapy (CVT). Results indicated that the maximum phonation time was significantly elongated in PVT(3.85±2.81s) but not in CVT(0.46 ± 2.24s), and the formant-distance was significantly increased in PVT(95.80 ± 112.71Hz) compare to CVT(-47.10 ± 84.28Hz). Our results indicated that by demanding appropriate amount of postural maintenance during vocalization, it might facilitate the recovery of speech-related functions. This study warranted larger-scale clinical trials to understand the impact, limitation, and optimization of posture-voice therapy for HD-PD.

## 1 Introduction

Hypokinetic dysarthria (HD) is a speech disorder unique to patients with Parkinson’s disease (PD)(Pinto et al., 2004). HD-PD traumatizes the overall quality of life with PD due to impaired communication and social participation(Miller et al., 2006), which tend to gradually deteriorate to the point of complete refusal to communicate(Schalling et al., 2017). Although both levodopa drugs and deep brain stimulation (DBS) have been proven to improve motor symptoms in patients with PD, neither has shown significant therapeutic effects on speech-related impairments(Tykalová et al., 2015; Hariz and Blomstedt, 2022).

Successful production of speech depends on a coordinated interplay among respiration, phonation, articulation, resonance, prosody, etc.(Yorkston, 1996). Therefore, direct training on these activities have been explicitly studied and administered in rehabilitation clinic for HD-PD(Herd et al., 2012). For example, phonation was instrumental in LSVT LOUD(Ramig et al., 2018), a widely tested and accepted therapeutic regime(Mahler et al., 2015). Respiration is emphasized in Respiratory Muscle Strength Training (RMST), where improvement in the patient’s respiratory muscle strength may result in a greater flow of air to stimulate voice production(Zhuang and Jia, 2022). Prosody is a key component of the SPEAK-OUT training program, thereby improving monopitch in patients with PD(Boutsen et al., 2023). In addition, posture and movement were also considered important in the selection of therapy goals for the elicitation of better voicing(Behrman, 2005). However, most of the existing therapies imposed minimal, if not none, requirements on how postures should be maintained during vocalization.

Incorrect posture and body movements could hinder the recovery of vocalization and even the overall axial symptoms (Angsuwarangsee and Morrison, 2002; Yoshii et al., 2016). In specific, a posture with improper alignment of the body tends to facilitate the voice production, due to the excessive the energy expenditure and strain on tissues(Arboleda and Frederick, 2008; Cardoso et al., 2019). Another postural abnormality found commonly with PD is with neck flexion(Yoshii et al., 2016). The soft tissue of the throat and the muscles that raise the larynx can alter due to imbalances in the neck and head, which can affect voice control and resonance (Cardoso et al., 2019). Therefore, therapeutic exercise with postural correction may provide addition benefits for voice production in PD. For instance, Yijinjing is a type of traditional Chinese exercise with explicit emphasis on the coordination of posture, which has been proven to alleviate joint pain(Cheng et al., 2022), increase muscle strength, and improve balance(Huang et al., 2022). The ninth method of Yijinjing – “Wohu Pushi (a crouching tiger to pounce for food)” – describes a posture-voice exercise. The Wohu Pushi method asks the trainee to imitate a crouching tiger by assuming a *lunging-and-clawing* posture, and it trains the voice production by imitation of a tiger’s roaring(Yan, 2023).

In this study, we added posture maintenance to a conventional voice therapy conforming with LSVT LOUD principles. We present a protocol of posture-voice therapy that PD patients with mild to moderate motor impairments could still participate. Using this protocol, we also examined the posture-voice therapy on speech function in PD patients who received deep-brain stimulation in subthalamic nuclei (STN-DBS). The posture-voice therapy was tested in comparison with the conventional voice therapy, both of which incorporated similar voice exercises but they differed in requirements on posture. Objective measures on acoustic and speech performances were acquired and analyzed. Our main hypothesis was that the posture-voice therapy would incur discernable short-term changes after one session of training. If proven, this study would provide support for a series of experiments to identify the longer-term clinical benefits of posture-voice therapy as well as its neurological mechanisms.

## 2 Methods

We designed a prospective study to compare speech improvement before and after training in patients with conventional voice therapy (CVT) or posture-voice therapy (PVT). The CVT was derived following LSVT LOUD principles with essential adaptations (Fig.1A), which ensured that the repetition, content, and perceived effort of voice training were similar between groups. We modified the conventional therapy inspired from the ninth method “Wohu Pushi” of Yijinjing, a traditional Chinese exercise (Fig. 1B). By requiring a *lunging-and-clawing* posture during the conventional voice therapy, the participants received posture-voice therapy in the PVT group (Fig. 1C). Details of CVT and PVT are specified below.

**Figure. 1.**
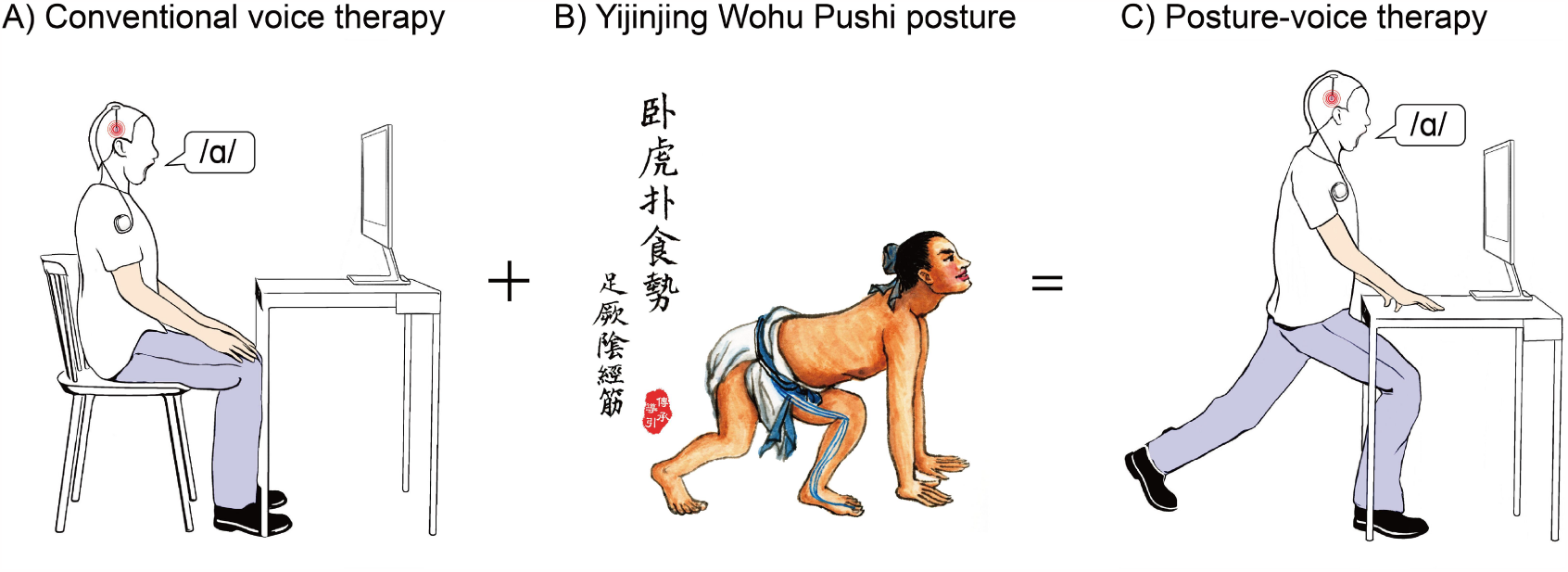
Illustration of the conventional voice therapy and the adapted posture-voice therapy. A) The body configuration in CVT group, the vocalization was performed while sitting in a chair. B) According to the ninth method of Yijinjing, the Wohu Pushi posture imitates a crouching tiger poised to pounce for food(Yan, 2023), which inspired the posture-voice therapy proposed in this study. C) The body configuration in PVT group, the participant assumes a leg-lunging and hand-clawing posture during vocalization.

### 2.1 Training protocols

In both groups, participants were required to accomplish 3 tasks of phonation specified below.

Tone-up (TU): The vowel /a/ is pronounced from low pitch to high pitch, with the entire pronunciation process lasting more than 10 seconds and maintaining a slight hold at the highest pitch.

Maximum sustained movements (MS): Prolong the vowel /a/ as much as possible while maintaining vowel pronunciation at 70% of your maximum SPL.

Tone-down (TD): The vowel /a/ is pronounced from high pitch to low pitch, with the entire pronunciation process lasting more than 10 seconds and maintaining a slight hold at the lowest pitch.

The primary differences between groups were with the posture maintenance during phonation tasks.

The CVT group, the participant performed the above phonation tasks while remaining seated in the chair throughout the vocalization training.

In the PVT group, the key requirement was the maintenance of a *lunging-and-clawing* posture during voice production. The lunging required that the participant started with a stance at shoulder-width, and lunged to keep the forward thigh horizontal. The forward knee should align with the toe. The behind leg should be extended straight. Meanwhile the trunk should stay upright.

The clawing required that the participant started with their hands shoulder-width apart, fingers spread, with the fingertips pressing against the tabletop. The elbows should be straight, shoulders relaxed, and chest raised. During vocalization, the cervical spine should be extended backward and the mouth opened as wide as possible. The experimenter helped the participant adjust the height of the table to the lowest clearance without losing stability.

The sequence of trials in both groups are listed in (Fig. 2). Each participant, either in the PVT or CVT group, accomplished 48 repetitions of vocalization tasks. Verbal feedback and encouragement were given to the participant after each trial of vocalization. Rests were given ad-hoc between trials as required by the participants.

**Figure. 2.**
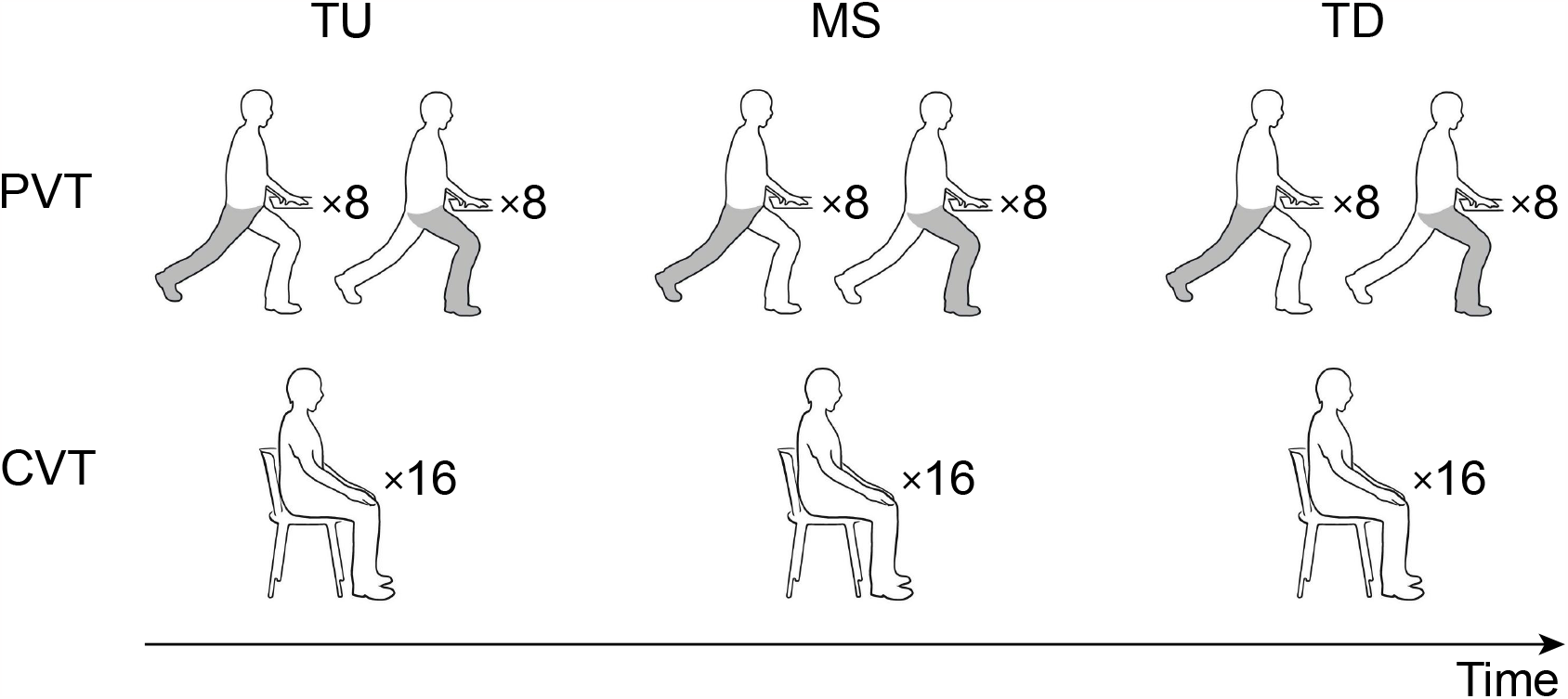
The sequence of the three tasks—Tone-up (TU), Maximum sustained movements (MS), and Tone-down (TD)—introduced in the training for two groups (PVT vs. CVT). The PVT group performs the exercises in the “Wohu Pushi” posture, with alternating leg configurations: eight repetitions with the right leg in the back, followed by eight repetitions with the right leg in the front. The CVT group performed each task in a seated position, with 16 repetitions of each task.

### 2.2 Study Design

Participants were recruited from February to June 2023 from the outpatient Department of Neurosurgery, Center for Functional Neurosurgery, Ruijin Hospital, Shanghai Jiao Tong University School of Medicine, Shanghai, China. The inclusion criteria were: (1) More than 6 months after bilateral STN-DBS surgery for PD; (2) No adjustments to medication or DBS stimulation parameters in the past 3 months; (3) VHI-10>11 (Arffa et al., 2012); (4) Within Stages I to IV on the Hoehn Yahr scale(Hoehn and Yahr, 2001); (5) No significant cognitive impairment (MMSE⩾25)(Folstein et al., 1975); (6) Age 18 and above; (7) Native Chinese speaker. The exclusion criteria were: (1) Diagnosed previously or currently with diseases that affect swallowing function (such as vocal nodules, gastroesophageal reflux, laryngeal cancer, and other laryngeal diseases); (2) Having difficulty breathing, fever, lung infection or chronic lung disease, or abnormal oral structure; (3) Due to various other reasons, the training tasks required for this study cannot be completed.

Allocation of participants into groups was accomplished using a computer program with random number generation (Fig. 3A). All PD participants were on levodopa medication with daily routine of dosage. Each patient underwent a speech assessment, followed by the phonation exercise as required by the allocated group. Immediately after training, another speech assessment was carried out. Each participant practiced 5-10 trials of phonation (either TD/MS/TU) as a familiarization to the protocol. The entire visit took approximately 1.5 hours (Fig. 3B). All participants gave written consent as approved by the Ethics Committee of Ruijin Hospital, School of Medicine, Shanghai Jiao Tong University (No. 224 of 2022). The training was administered by certified inheritors of Yijinjing (The National List of Intangible Cultural Heritage of China, Project No. IX-2) and licensed LSVT-LOUD therapists (ONLOUDCH1121-52, 0822-18, 0822-19, 1122-04).

**Figure. 3.**
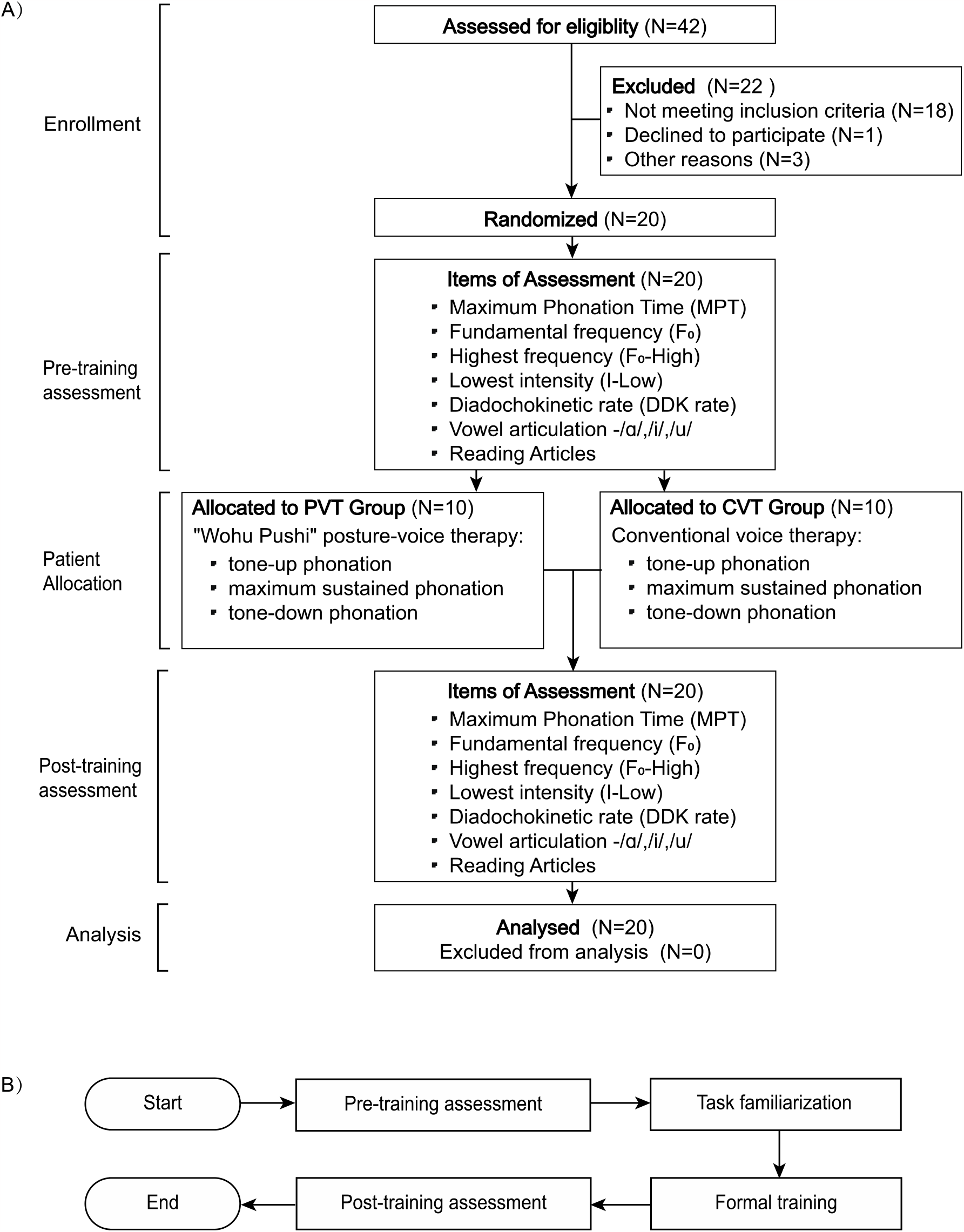
Flowchart of the trial. A) the flow diagram for recruitment, group allocation, assessment, and analysis. B) procedure of each visit: participants received interventions of voice training. Data were analyzed before and after the intervention.

### 2.3 Data Acquisition

Speech recordings were performed in a quiet room with a low ambient noise level using a condenser microphone placed approximately 30 cm from the subject’s mouth (Fig. 4). Speech signals were sampled at 48 kHz with 16-bit resolution. Our acoustic indicators including MPT, DSI, SPL/a/, SPL”Reading”, DDK rate, and vowel production(/a/,/i/,/u/), collected through the following acoustic tasks(Wuyts et al., 2000; Karlsson and Hartelius, 2019; Rusz et al., 2021; Boogers et al., 2022).

**Figure. 4.** Actual scenes of therapeutic training and assessments. A) A participant performing “Wohu Pushi” posture-voice therapy. The participant performed vocal exercises by leaning with both hands on a height-adjustable table, maintaining a *lunging-and-clawing* posture. B) the acoustic assessment of participants using lingWAVES. Participants vocalized facing a microphone, while an experimentalist recorded the acoustic parameters of voice.

Maximum phonation time (MPT, in seconds) – take a deep breath, and sustain the vowel “ah” as long as possible, record the longest phonation out of three measures as MPT;

Fundamental frequency (F_0_, in Hz) - maintain a comfortable volume and a consistent tone while sustaining the vowel sound /a/ for more than 3 seconds;

Highest frequency (F_0_-High, in Hz) - gradually increase the tone of “ah” from normal speaking to the highest point and sustain it for 1-2 seconds;

Lowest intensity (I-Low, in dB) - Start at a normal volume, sound “ah”, and gradually decrease the volume;

Diadochokinetic rate (DDK rate, in syllables per second) - produce the syllables /pa/, /ta/, and /ka/ as quickly as possible for as long as you can;

Vowel production - pronounce the vowels /a/, /i/, and /u/ as smoothly and comfortably as possible, maintaining the sound for more than 3 seconds;

SPL/a/ (in dB) - acoustic record reference fundamental frequency;

SPL”Reading” (in dB) - read the story of “The North Wind and the Sun” at a comfortable pitch and loudness, as if you are having a regular conversation.

(Figure. 4 is omitted due to exposure of patient images; the figure is available upon request to the corresponding authors.)

Using the PRAAT software, select the formant one frequency F1 and the formant two frequency F2 for the vowels /a/, /i/, and /u/ respectively. The software LingWAVES (Software Version 3.2, Wevosys) was used for voice measurements. Analyze the acoustic data of the participants using PRAAT (version 6.3.08) software.

### 2.4 Outcome measures and statistical analysis

The primary outcome measure in this study was Δ MPT, defined as the difference in maximum phonation time before and after the training.

Another outcome measure was the formant-distance, defined as the frequency difference between the formant one of /a/ and /*i* /, i.e. Δ*F*1 = *F*1/a/− *F*1/*i* / (Mautner, 2016).

The third outcome measure was the vector-similarity between the participant-pronounced vector and the group-averaged vector, which was calculated based on the one and two formants of each vowel. Since the first two formants of vowel formed a 2-dimensional vector on the frequency plane (Niu et al., 2015), we were interested in the similarity between the participant-pronounced vector and the group-averaged vector. This outcome measure aimed to quantify whether a participant showed congruent changes compared to his/her group trend. It was calculated as follows:

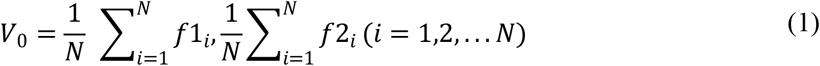

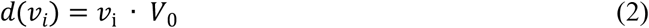

where *V*_0_ denotes the averaged formant-vector across all participants within a group, and *d*(*v*_*i*_) denotes the inner product between an individual vector and the group-averaged vector.

Statistical analyses were conducted using R (version 4.3.0). Between-group comparisons were conducted either using Welch’s t-test, or by a 2 × 2 two-way repeated-measure analysis of variance (RM-ANOVA) with factors of GROUP (PVT vs. CVT) and EVALUATION (before training, after training) formulated as follows:

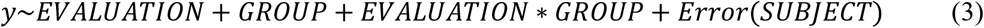

## 3 Results

### 3.1 Baseline characteristics of the participants

We screened 42 candidates who agreed to participate. A total of 20 participants finished the study, with 10 in the PVT group (3 male and 7 females, age 62.3 ± 9.29) and 10 in the CVT group (6 male and 4 females, age 65.9 ± 5.42). No statistical differences were found between groups in age, sex, months since diagnosis, months since surgery, Hoehn-Yahr (HY) scale, Unified Parkinson’s Disease Rating Scale III (UPDRS III), or Voice Handicap Index-10 (VHI-10) (Table 1). Refer to the supplementary materials for details about the stimulation setting.

**Table 1.**
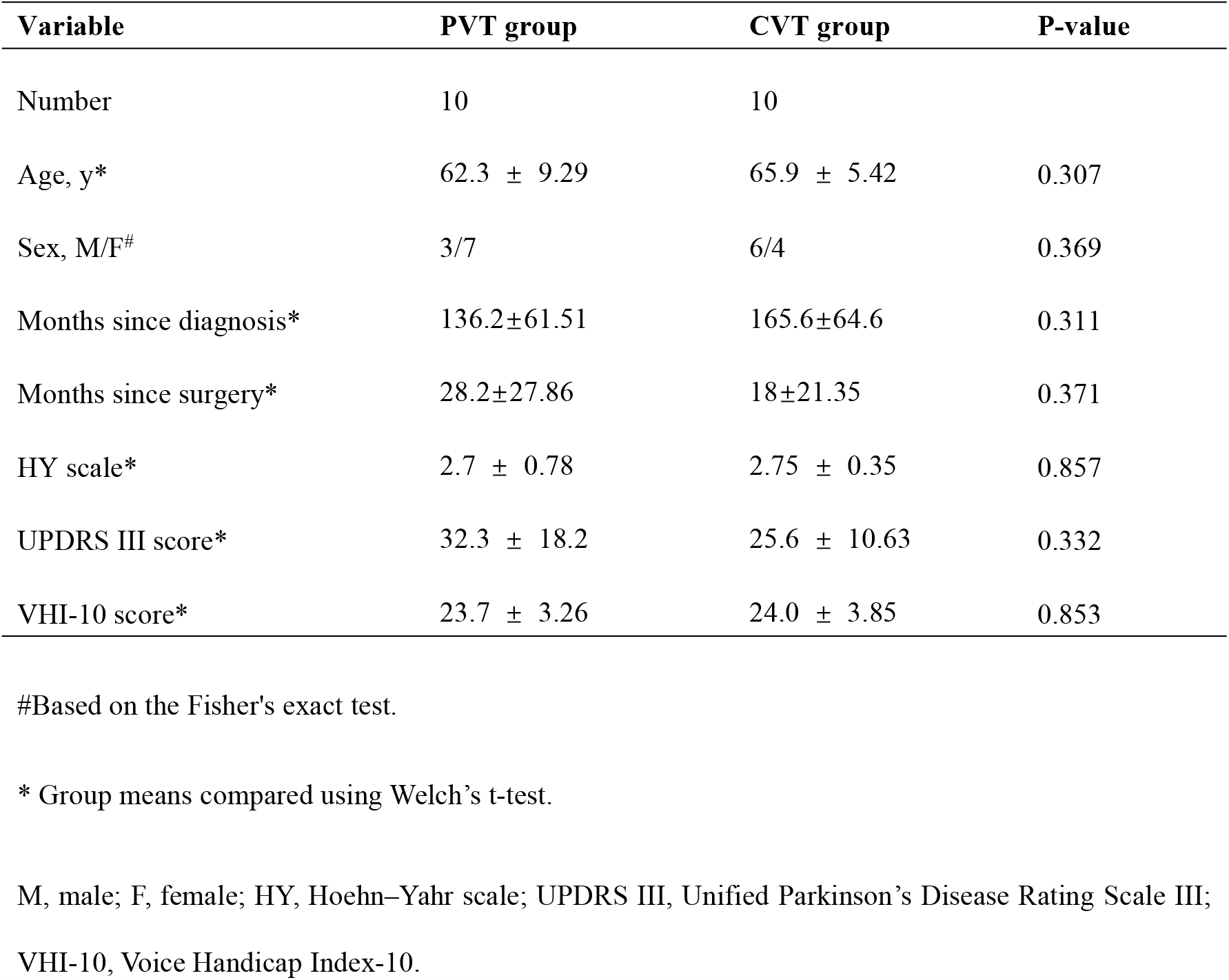
Baseline characteristics of the participants.

| Variable | PVT group | CVT group | P-value |
| --- | --- | --- | --- |
| Number | 10 | 10 |  |
| Age, y* | 62.3 ± 9.29 | 65.9 ± 5.42 | 0.307 |
| Sex, M/F<sup>#</sup> | 3/7 | 6/4 | 0.369 |
| Months since diagnosis* | 136.2±61.51 | 165.6±64.6 | 0.311 |
| Months since surgery* | 28.2±27.86 | 18±21.35 | 0.371 |
| HY scale* | 2.7 ± 0.78 | 2.75 ± 0.35 | 0.857 |
| UPDRS III score* | 32.3 ± 18.2 | 25.6 ± 10.63 | 0.332 |
| VHI-10 score* | 23.7 ± 3.26 | 24.0 ± 3.85 | 0.853 |

<sup>#</sup>Based on the Fisher's exact test.
\* Group means compared using Welch's t-test.

M, male; F, female; HY, Hoehn–Yahr scale; UPDRS III, Unified Parkinson’s Disease Rating Scale III; VHI-10, Voice Handicap Index-10.

### 3.2 Maximum phonation time

Voice time series of /a/ pronunciation from representative participants in each group are shown in Fig.5. As can be seen, Subject09 in the PVT group performed with an increase in MPT after training, while Subject107 in the CVT group also had an increase in MPT, but the change was smaller. Pooled data suggest that the PVT group exhibited an overall increasing trend in MPT scores post-intervention (Fig. 6A), while the CVT group showed inconsistent MPT changes among participants (Fig. 6B). A 2×2 two-way repeated-measures ANOVA (Fig. 6C) indicated a significant main effect of EVALUATION on MPT (F_1,18_ =14.305, p<0.01). No significant main effect for GROUP (F_1,18_ = 0.064, p = 0.803) was found. The interaction between GROUP and EVALUATION was significant (F_1,18_ =8.843, p<0.01), suggesting that the PVT and CVT groups differed in MPT score changes over time. Post-hoc comparison with the Tukey test showed a significant increase in MPT for the PVT group from “Before” to “After” (p < 0.001), but the changes were non-significant in the CVT group (p = 0.939).

**Figure. 5.**
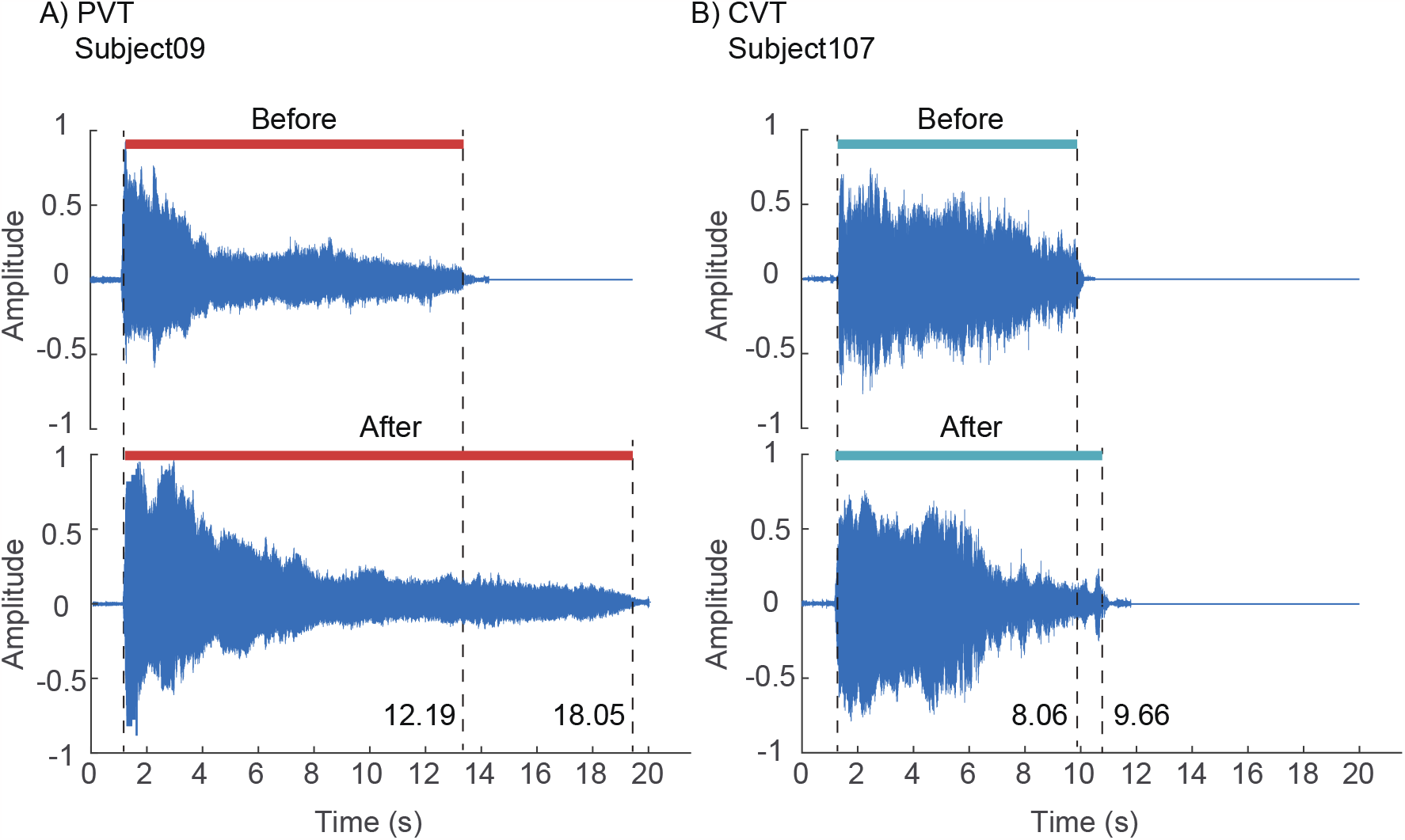
Comparison of MPT duration between two subjects. A) There was a representative participant (Subject09) in the PVT group. The MPT before therapy was 12.19 seconds, which increased to 18.05 seconds after the therapy. B) There was a representative participant (Subject107) in the CVT group. The MPT before therapy was 8.06 seconds, which increased to 9.66 seconds. The increase in the two participants was 48% and 19%, respectively.

**Figure. 6.**
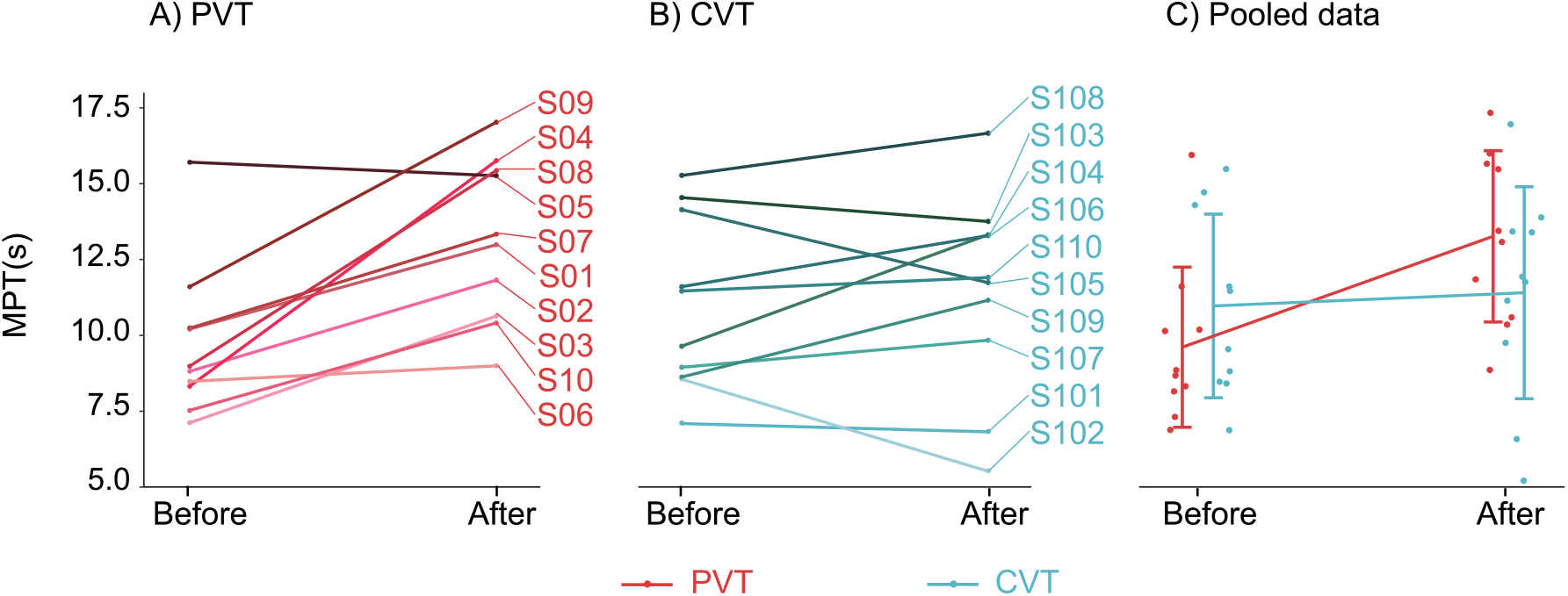
Comparison of MPT duration between PVT and CVT groups. A) Changes in MPT duration before and after training in PVT Group. red lines indicate the MPT duration change for each patient. B) Changes in MPT duration Before and After training in CVT Group. blue lines represent the MPT duration change for each patient. C) Comparative analysis of the mean slope changes in MPT duration. MPT increased more in the PVT group as compared to the CVT group.

### 3.3 Vowel production

We plotted the vector change graph on a two-dimensional plane, with the starting and ending points of the vector being the formant frequency before and after training, respectively(Fig. 7)(Niu et al., 2015). Pooled data suggest that the PVT group exhibited an overall increasing trend in formant-distance (ΔF1) after the therapy (Fig.8A), while the majority of participants in CVT group showed a decline (Fig.8B). A 2×2 two-way repeated-measures ANOVA (Fig. 6C) indicated no significant main effect of GROUP on ΔF1 (F_1,18_ = 2.856, p = 0.108) or main effect of EVALUATION (F_1,18_ = 1.436, p = 0.246). However, the interaction between EVALUATION and GROUP was significant (F_1,18_ = 12.362, p < 0.01). Post-hoc comparison with the Tukey test showed a significant increase in ΔF1 within the PVT group (p <0.05) but not in CVT group (p = 0.383).

**Figure. 7.**
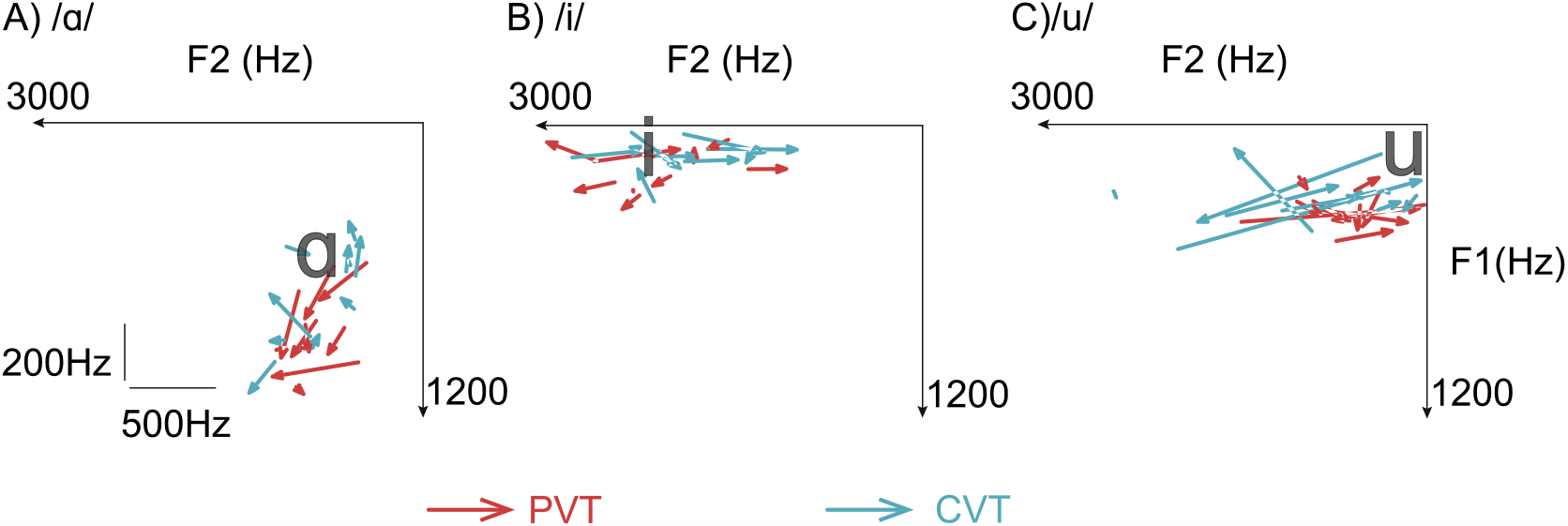
The formants one and two for vowels produced by all participants. Changes in vowel production are depicted as vectors pointing from before the training to after the training, either for /a/ in panel A), /i/ in panel B) or /u/ in panel C). The black bold symbols represent the average formant frequencies (Hz) for U.S. English vowels produced by 45 males (Hillenbrand et al., 1995).

**Figure. 8.**
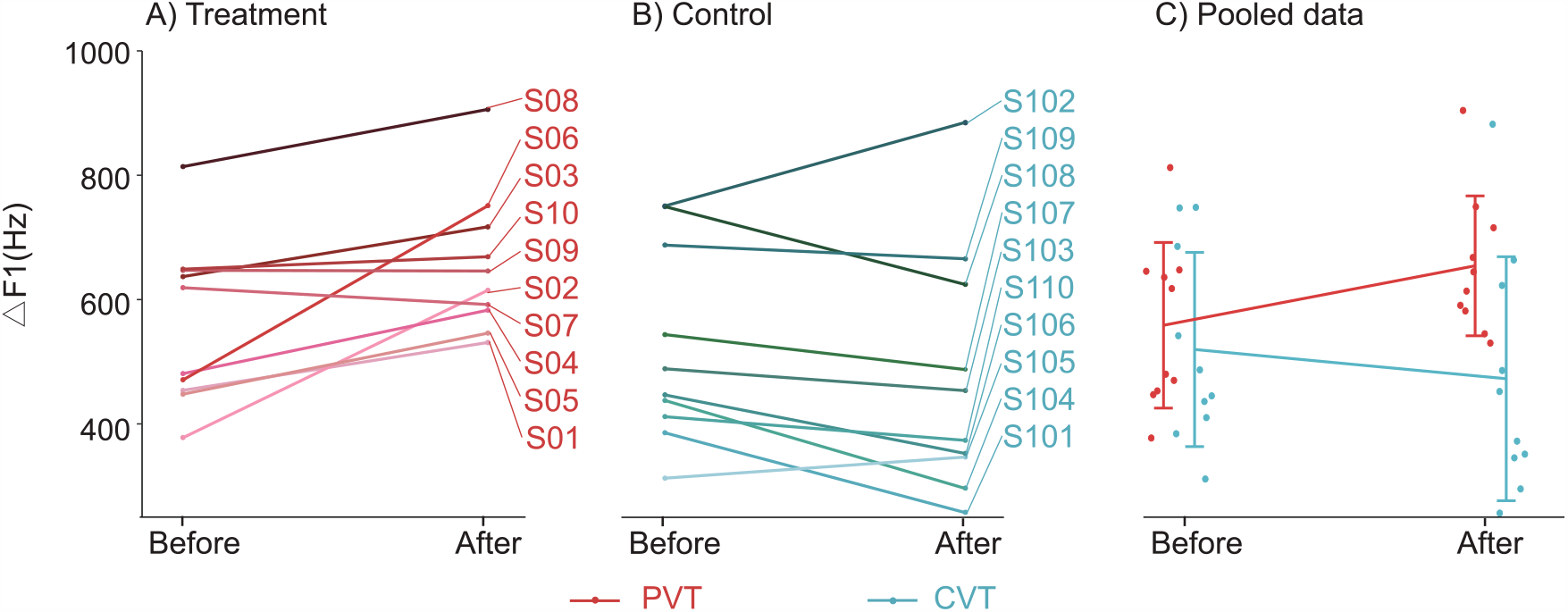
Comparison of ΔF1 between PVT and CVT Groups. A) Changes in ΔF1 before and after training in the PVT Group, with each red line segment representing the ΔF1 for a participant. B) Changes in ΔF1 before and after training in the CVT Group, with each blue line segment indicating the ΔF1 for a participant. C) Pooled data of ΔF1, the PVT group showed an increasing trend, while the CVT group showed a decrease after training.

The vector-similarity *d*(*v*_*i*_) between the individual vectors and group-averaged vectors wasanalyzed. Welch’s t-test showed that for the vowel /a/, the vector-similarity in PVT group was larger compared to that in CVT group (t_9.842_ = 3.165, p <0.05, Fig. 9A); Welch’s t-test was chosen because it allowed for comparisons between groups that did not confirm homoscedasticity. For the vowel /i/, the vector-similarity was smaller in PVT compared to CVT (t_9.181_=-2.691, p<0.05, Fig. 9B). Vowel /u/ showed no significant changes between groups (t_18_=0.857, p=0.402, Fig. 9C).

**Figure. 9.**
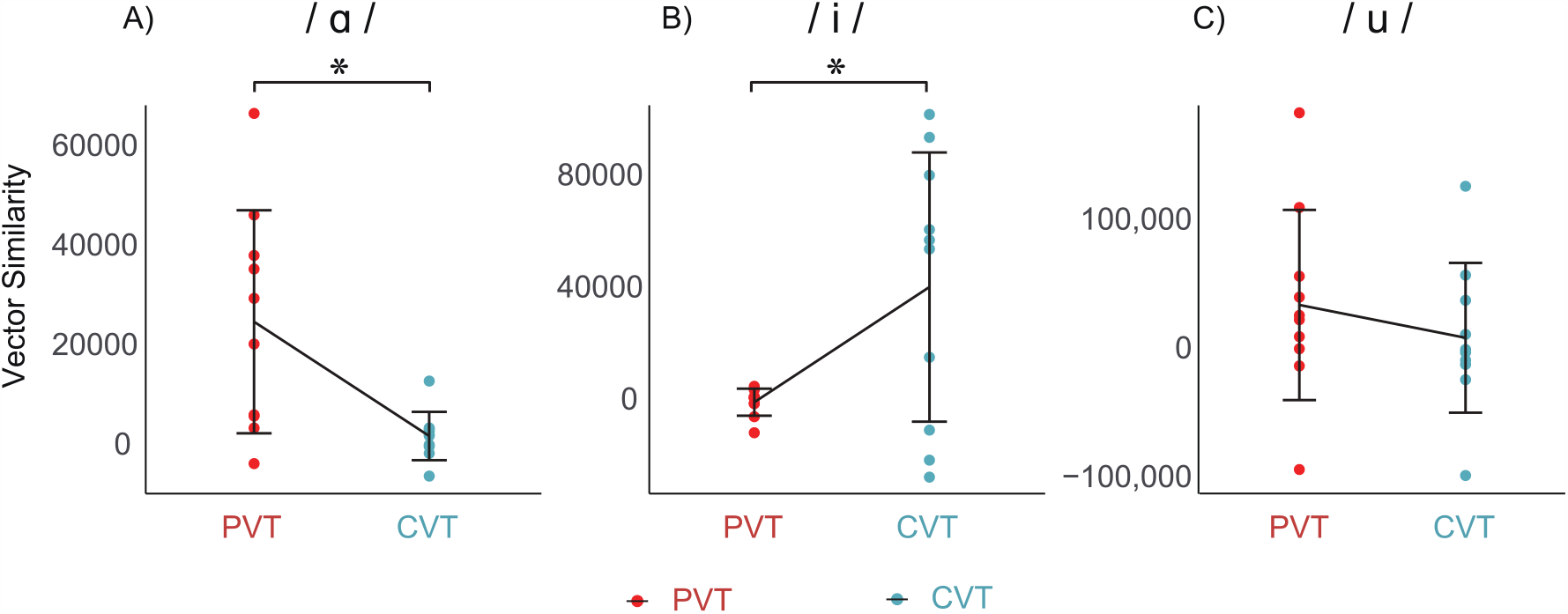
Vector similarities for vowel production in two groups. A) the results for the vowel sound /a/ in both groups. Significant differences were identified, indicating that the average group trend in PVT group is much stronger than CVT group. B) The vector similarity for the vowel /i/, a significant difference existed between the two groups. C) No significant difference between groups in terms of vector similarity for the vowel /u/.

### 3.4 DSI, SPL, DDK rate

For other acoustic measures, we did not detect any significant main effects of GROUP or EVALUATION (Fig.10), and no significant interactions were found.

**Figure. 10.**
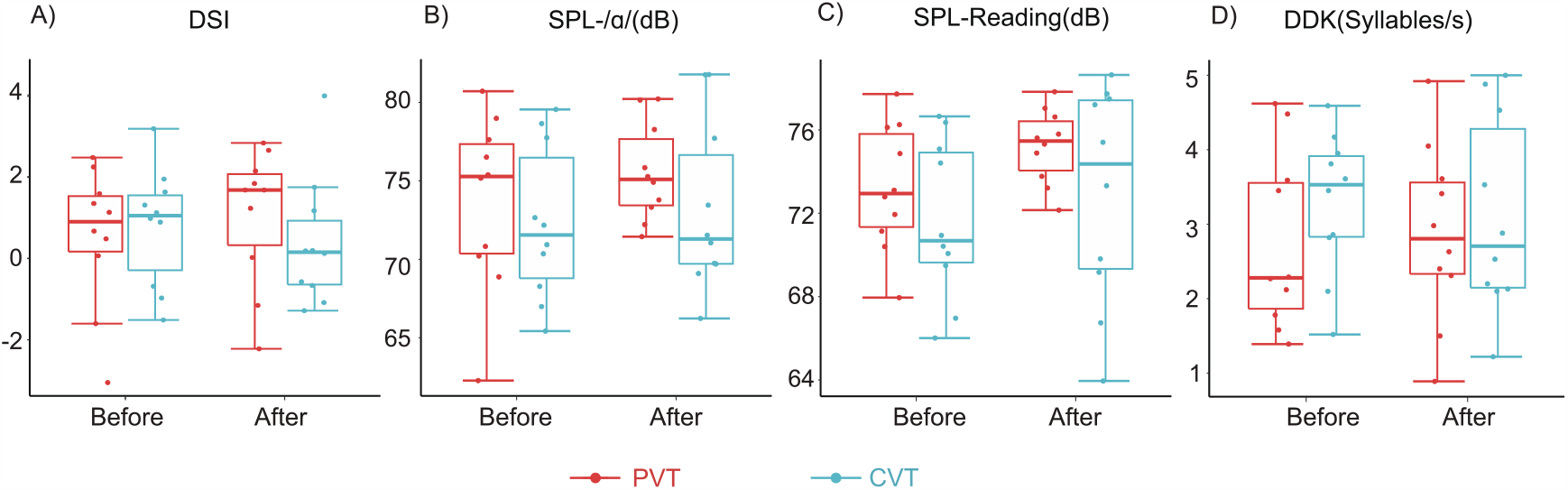
Four acoustic measures between PVT group and CVT group. A) dysphonia severity index rate (DSI), B) the vowel sound / a /, C) Reading articles, and D) diadochokinetic rate (DDK). No significant effects of GROUP or EVALUATION were found on these acoustic measures.

### 3.5 Correlation betweenΔMPT and other factors

Due to the significant improvement shown by MPT in the results, we continued to explore whether changes in MPT (ΔMPT) could be explained by known clinical profiles of participants. Linear regressions showed a significant correlation between ΔMPT and the months-since-diagnosis, i.e. the longer months since diagnosis, the more significant the improvement in MPT after one training session. The linear relationship is y = 0.026x + 0.372 (one-tailed p<0.05, R^2^ = 0.311, Fig. 11). ΔMPT was not significantly correlated with UPDRS III or VHI-10.

**Figure. 11.**
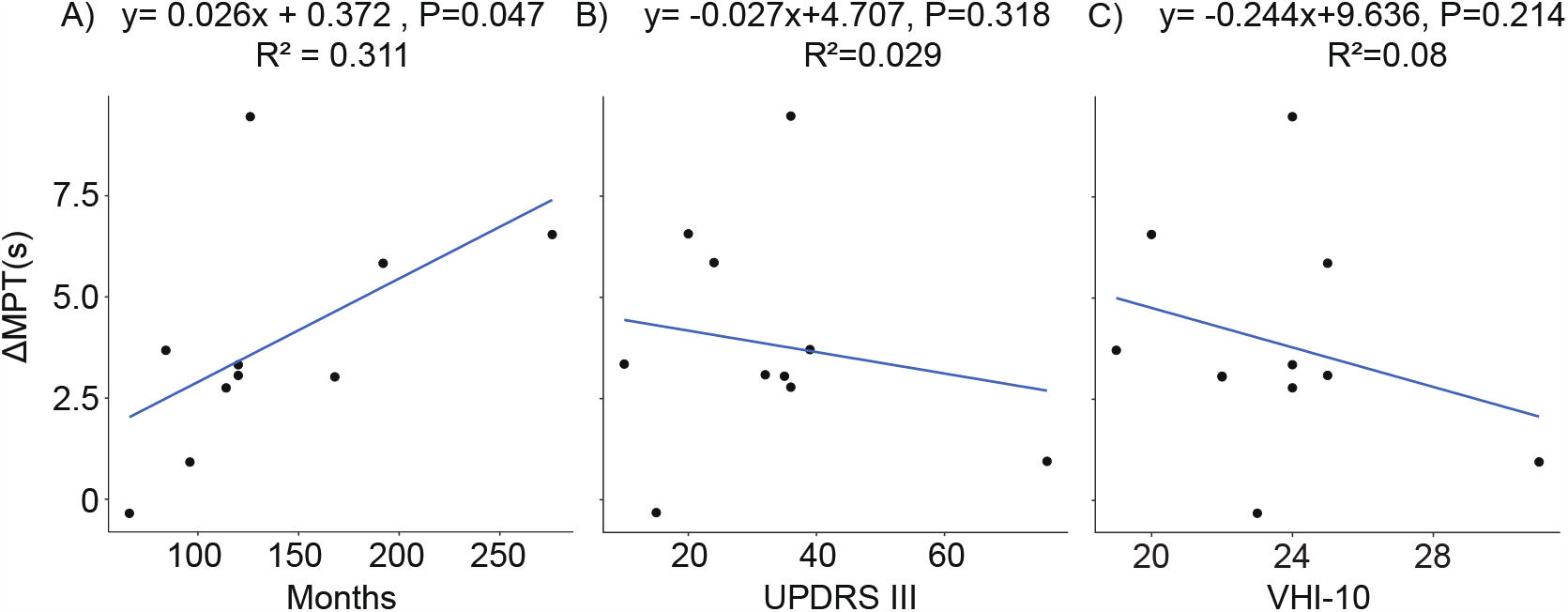
Linear regression of ΔMPT and clinical factors. The linear regression exploring the relationship between ΔMPT and months since diagnosis showed a significant correlation (p < 0.05, panel A). The linear relationship was non-significant between ΔMPT and UPDRS? (B) or VHI-10 (C).

## 4 Discussion

This study developed a protocol for conducting voice training for PD patients after STN-DBS. The protocol specified that a *lunging-and-clawing* posture should be maintained during vocalization. A pilot study was conducted to examine the short-term benefit of posture-voice therapy compared with conventional voice therapy that did not impose posture requirements. Results showed that after one session of training, the posture-voice therapy (PVT) managed to elongate the maximum phonation time and the formant-distance better than the conventional voice therapy (CVT). Our results suggested more prominent improvements if the patients received longer periods of “Wohu Pushi” posture-voice therapy. This short-term protocol was not sensitive enough to detect significant changes in acoustic measures including DSI, SPL, and DDK rate. Overall, our results indicated that adding posture requirements to conventional voice therapy for PD patients could facilitate recovery on voice and speech production.

One reason why posture-voice therapy elongated MPT more than did the conventional voice therapy, is that MPT associates with the respiratory capacity during speech production(Zhuang and Jia, 2022). By assuming the typical “Wohu Pushi”posture, subjects may have learned to inspire to a higher percentage of their vital capacities, due to the stretching of the respiratory muscles (including the intercostal muscles, the sternocleidomastoid muscles and pectoralis major muscles), thereby increasing the amount of available air stream used for phonation.

It is noteworthy that the formant-distance in PVT group was larger than that in CVT group, even though both groups emphasized on keeping the mouth open during training. Formant-distance is a quantitative measure of the acoustic form of the degree of mouth opening, which is generally proportional to the quality of the pronunciation(Sapir et al., 2011; Ikui et al., 2015). The repeated flexion and extension of the cervical spine during training in the PVT group may stimulate the proprioception of the relevant muscles in the jaw, thereby improving the sensory processing of the jaw.

Our current experimental setup did not allow us to differentiate the contribution between leg lunging and hand supporting. Although this study demonstrates that overall postural vocalization can improve certain acoustic indicators, whether the upper or lower limbs contribute more to vocalization, allowing us to optimize it more in the future, remains to be further investigated.

In conclusion, this pilot study suggests that by requiring adequate posture maintenance in a conventional voice therapy, we have developed a clinically viable protocol that PD patients with STN-DBS could accomplish even with mild to moderate motor impairments. The posture-voice therapy is likely more effective in improving MPT and formant-distance compared to the conventional voice therapy.

## Abbreviations

CVT: conventional voice therapy
DBS: deep brain stimulation
DDK: diadochokinetic
DSI: dysphonia severity index
HD: hypokinetic dysarthria
HY: Hoehn-Yahr scale
LSVT: Lee Silverman Voice Therapy
MMSE: mini-mental state examination
MPT: maximum phonation time
MS: maximum sustained movements
PD: Parkinson’s disease
PVT: posture-voice therapy
RMST: Respiratory Muscle Strength Training
RM-ANOVA: repeated-measures analysis of variance
SPL: sound pressure level
STN: subthalamic nucleus
TD: tone-down
TU: tone-up
UPDRS: Unified Parkinson’s Disease Rating Scale
VHI: Voice Handicap Index

## Author Contributions

Conceptualization, C.M.N. and D.Y.L.; formal analysis, X.S. and Y.X.S; funding acquisition, D.Y.L. and C.M.N.; investigation, X.S. and J.Y.; methodology, X.S., M.Y., J.Y., and C.M.N.; project administration, D.Y.L. and C.M.N.; resources, D.Y.L.; supervision, S.Q.Y. and Y.W.; writing—original draft, X.S.; writing—review and editing, L.B.W, D.Y.L., and C.M.N. All authors have read and agreed to the published version of the manuscript.

## Institutional Review Board Statement

This study was approved by the Ethics Committee of Ruijin Hospital, School of Medicine, Shanghai Jiao Tong University (no. 224 of 2022).

## Informed Consent Statement

Informed consent was obtained from all subjects involved in the study.

## Data Availability Statement

The data presented in this study are available on request from the corresponding author.

## Acknowledgments

The authors thank Ms. Lu Xu for her assistance with patient recruitment and baseline assessment. We also thank Mr. Qiang Xu from RJH-FFTAI Joint Laboratory for development of essential software to facilitate the study.

